# Cannabinoid type 1 receptor availability imaging in attention-deficit/hyperactivity disorder using [^18^F]MK-9470 positron emission tomography: study protocol of the CB1-ADHD study

**DOI:** 10.64898/2026.01.07.25342349

**Authors:** Isabelle Miederer, Marina Louise Schofer, Christian Ruckes, Wolfgang Retz, Mathias Schreckenberger, Alexandra Sebastian

**Author notes:** Correspondence: Isabelle Miederer.

## Abstract

**Background:** Attention deficit/hyperactivity disorder (ADHD) is a neurodevelopmental disorder characterized by attentional deficits, hyperactivity and impulsivity that often persists into adulthood. While dysfunction of dopaminergic neurotransmitter systems has been observed, underlying mechanisms remain incompletely understood. Current treatments with methylphenidate and amphetamines show limited long-term effectiveness and do not address broader clinical needs. The endocannabinoid system represents a promising therapeutic target. Cannabinoid type 1 (CB1) receptors are highly concentrated in the prefrontal cortex and striatum, brain regions central to ADHD pathophysiology. The "dual pathway model of ADHD" describes deficits in inhibition-related executive functions and reward-related functions, both mediated by fronto-striatal networks. Striatal CB1 receptor availability correlates negatively with impulsivity. Given the interaction between dopaminergic and endocannabinoid systems, CB1 receptors may play a key role in ADHD pathogenesis.

**Methods:** This controlled study will investigate CB1 receptor availability in ADHD using positron emission tomography (PET) with the CB1-selective radiotracer [^18^F]MK-9470. The primary outcome is the CB1 receptor distribution volume (V_T_) in the striatum. Secondary outcomes include CB1 receptor availability in prefrontal cortex and other ADHD-relevant brain regions, plasma concentrations of endocannabinoids (anandamide, 2-arachidonoylglycerol), and validated neuropsychological assessments of attention and impulsivity. We will compare three groups (n=34 each): medication-naïve participants with ADHD, methylphenidate-treated participants with ADHD, and healthy controls. Statistical analysis will employ ANOVA with post-hoc comparisons and correlation analyses between neuroimaging and behavioral measures.

**Discussion:** This first investigation of the endocannabinoid system in ADHD will provide crucial insights into disease mechanisms and identify potential therapeutic targets. Results may inform development of novel cannabinoid-based treatments and improve evidence-based therapeutic strategies for ADHD management.

**Trial registration**: German Clinical Trials Register (DRKS-ID: DRKS00037526, Registration date: 25 September 2025)

## Introduction

### Background and rationale

Attention-deficit/hyperactivity disorder (ADHD) is a neurodevelopmental disorder that often persists into adulthood. It is characterised by core symptoms of attentional deficit, hyperactivity and impulsivity; common associated symptoms in adults are emotional instability and disorganised behaviour (1). In Germany, the estimated point prevalence rates are 4-5 % in children and 2-3 % in adults. Males are diagnosed more frequently than females (2-3:1 in children and adolescents, 1.5-2:1 in adults). ADHD is associated with comorbid disorders such as depression, anxiety, personality disorders, addictive disorders, eating disorders and restless legs syndrome. It is also associated with psychosocial impairment and health risks, such as a high risk of accidents, relationship problems, occupational difficulties, antisocial behavior and delinquency, resulting in a substantial economic burden (2–4). According to the Global Burden of Diseases, Injuries and Risk Factors Study published by the World Health Organization, the category ’other mental and substance use disorders’, which includes ADHD, is one of the 20 leading causes of years lived with disability in 2016 (5).

Increased impulsivity is a central feature of ADHD (6). Prominent models, such as the "dual pathway model of ADHD" (7,8), suggest that individuals with ADHD have deficits particularly in two facets of impulsivity, namely inhibited-related executive functions (e.g. motor inhibition) and/or in reward-related functions (e.g. delay discounting) (9). Neuroimaging studies have consistently implicated alterations in frontoparietal, dorsal frontostriatal and mesocorticolimbic circuits as well as in default mode and cognitive control networks to ADHD (for a review see (10)). In terms of neurotransmitter systems, dysfunctions in the dopaminergic and noradrenergic systems have been found in ADHD (for a review see (11)), and the serotonergic system is also receiving increasing attention (12). In addition, ADHD is increasingly understood as a network disorder in which not only region-specific deficits are present; rather, the connectivity of various brain networks is impaired, which is considered a neurobiological marker of ADHD (13–15). However, the neuronal mechanisms underlying ADHD are incompletely understood.

Treatment options for ADHD include psychotherapy and/or pharmacotherapy. According to the current S3 guideline "Attention-Deficit/Hyperactivity Disorder (ADHD) in Children, Adolescents and Adults" (12), the stimulants methylphenidate and amphetamine, which act on dopaminergic and noradrenergic neurotransmitter systems, are the drugs of first choice. Stimulant therapy is well established, and numerous randomised controlled clinical trials have shown positive short-term effects of stimulant treatment for ADHD symptoms in children and adults (3,16). The modulating effects of stimulants on the intrinsic connectivity of brain networks are also evident at the brain network level, despite methodological heterogeneity (17). However, the response rate is around 70 % and stimulants are contraindicated in some cases of adult ADHD because of sympathomimetic side effects such as increased blood pressure and pulse (3). There is therefore a high medical need for the further development of ADHD treatments.

A new and promising approach to better understand the underlying neural mechanisms of ADHD and to develop new therapeutic approaches is to investigate the involvement of the endocannabinoid system. The endocannabinoid system is composed of cannabinoid type 1 and 2 receptors, its endogenous ligands (endocannabinoids) (anandamide or N-arachidonoylethanolamine (AEA) and 2-arachidonoylglycerol (2-AG)) and its synthesizing and degrading enzymes. Other relevant biomarkers are the endocannabinoid-like compounds palmitoylethanolamide (PEA) and oleoylethanolamide (OEA) as well as arachidonic acid (AA) (for review see (18)). CB1 receptors are G-protein coupled receptors and found in high concentrations in striatum, prefrontal cortex and cerebellar cortex (19,20). These are brain regions that are involved in cognitive processes, such as attention, executive functions including impulse control, and motivation and reward and that are relevant to ADHD (10). CB1 receptors are mainly located presynaptically on excitatory and inhibitory neurons, such as glutamatergic, GABAergic, maybe dopaminergic, and others (21). Their endogenous ligands, the endocannabinoids, act as retrograde neurotransmitters, modulating the release of other neurotransmitters, such as glutamate or GABA, at the axon ends. The CB1 receptor has been described as being of great importance to many neurotransmitter systems for receptor signalling, changes in neurotransmitter release and neuronal firing (for review see (22)). In the dopaminergic system, including the striatum, the endocannabinoids AEA and 2-AG are abundant and modulate dopamine transmission (11). Several papers have shown that stimulation of CB1 receptors increased the firing rate of dopaminergic neurons and facilitated dopamine release in the striatum (for reviews see (11,23,24)). Conversely, there is increasing evidence that endocannabinoid signalling is controlled by the dopaminergic system, with D2 receptors controlling AEA production in the striatum and Gi/o protein availability to CB1 receptors, and controlling endocannabinoid-mediated functional synaptic plasticity of GABAergic neurons (25). Due to its ubiquitous distribution in the brain and modulation of neurotransmitter release, the endocannabinoid system is increasingly implicated in mental disorders, including ADHD (26). There are also indications from a genetic point of view. An investigation of the association between polymorphisms in the cannabinoid receptor 1 (CNR1) gene (rs806379 and rs1049353) and impulsivity-related phenotypes found that adversity early in life is associated with increased impulsivity in homozygous carriers of the rs806379 A and rs1049353 T alleles compared to homozygous carriers of the respective major allele (27). These findings support previous results linking CNR1 polymorphisms with impulsivity and associated psychiatric disorders such as substance dependence or ADHD; this suggests a genetically mediated effect on endocannabinoid signalling as the neurobiological basis of impulsive behaviour (27–32).

To date, ADHD has been linked to the dopamine system: striatal alterations have been demonstrated in ADHD (33), and stimulant treatment acting on the dopamine system has been shown to have therapeutic effects on central dysfunctions in ADHD, namely motor impulsivity and delay discounting (34–38). Importantly, the dopamine system is modulated by the endocannabinoid system (11,23,24) and there is evidence for the reverse (25). Given the dopamine hypothesis, the dopaminergic involvement in the striatum and in motor impulsivity and delay discounting in ADHD, and the interaction between the dopamine and endocannabinoid systems, it is conceivable that the endocannabinoid system may be involved in the aetiology and pathogenesis of ADHD.

## Objectives

PET studies have shown that CB1 receptor availability is reduced or increased in psychiatric disorders associated with impulsivity (39–50). These studies demonstrate that the endocannabinoid system exhibits different patterns (receptor up- and receptor downregulation) in different brain regions, which are considered as compensatory mechanisms for altered endogenous ligand concentrations. Receptor up- or downregulation could be explained, at least in part, by an (in-)sufficient compensatory mechanism to regulate an under- or overstimulated system (11). In high novelty seekers, CB1 receptor availability was globally reduced compared to low novelty seekers and negatively correlated in the striatum with the sub-dimension "extravagance", which is a measure of impulsivity (51). As high novelty seeking is one of the most important features of ADHD, we propose the following hypotheses: Primary hypothesis: Impulsive behaviour is associated with downregulation of CB1 receptors in the striatum (51). In addition, concentrations of the endocannabinoids AEA and 2-AG are increased in unmedicated participants with ADHD compared to healthy subjects (52), suggesting a compensatory downregulation of CB1 receptors (40,43,49–51). Therefore, our primary hypothesis is that the CB1 receptor availability will be lower in unmedicated participants with ADHD, primarily in the striatum, compared to healthy controls. Secondary hypotheses:

1. Methylphenidate effects on CB1 receptors: Methylphenidate is an approved drug for the treatment of ADHD in Germany, but its effect on the endocannabinoid system has not been studied. There is evidence that methylphenidate suppresses endocannabinoid concentrations in the brain (53). We assume that this leads to a compensatory upregulation of CB1 receptors, as this has been described as a possible response to altered endogenous ligand concentrations (40,43,47,49–51). Therefore, we hypothesize a) higher CB1 receptor availability in methylphenidate-treated participants with ADHD compared to unmedicated participants with ADHD, and b) lower CB1 receptor availability in methylphenidate-treated participants with ADHD compared to healthy controls.
2. Regional striatal differences: According to the "dual pathway model of ADHD", different aspects of impulsivity, i.e. inhibition-related (e.g. motor impulsivity) and reward-related (e.g. delay discounting), are mediated by dorsal and ventral striatal involvement respectively (8,33). Therefore, based on the above reasoning, we expect for both dorsal and ventral striatum a) lower CB1 receptor availability in unmedicated participants with ADHD compared to healthy controls, b) higher CB1 receptor availability in methylphenidate-treated participants with ADHD compared to unmedicated participants with ADHD, and c) lower CB1 receptor availability in methylphenidate-treated participants with ADHD compared to healthy controls.
3. Endocannabinoid concentrations: There is evidence that FAAH activity is reduced in the blood of individuals with ADHD compared with healthy controls (54), suggesting increased AEA levels. In addition, AEA and 2-AG have been shown to be elevated in untreated individuals with ADHD compared with healthy controls (52). There is also evidence that methylphenidate reduces levels of AEA and 2-AG (53). We therefore expect a) higher levels of AEA and 2-AG in unmedicated participants with ADHD compared to healthy controls, b) lower levels of AEA and 2-AG in participants with ADHD treated with methylphenidate compared with those not taking medication, and c) higher levels of AEA and 2-AG in - participants with ADHD treated with methylphenidate compared to healthy controls.
4. Behavioral measures: A large body of research has shown deficits in ADHD on various measures of impulsivity, such as the stop-signal task (55–57), delay discounting task (58,59), and questionnaires. Consequently, we expect to see a) higher self-reported impulsivity scores in unmedicated participants with ADHD compared to healthy controls, as well as more pronounced impulsivity in objective behavioural measures (namely longer stop-signal response time (SSRT) and steeper delay discounting, as indexed by a smaller area under the curve (AUC)), b) Lower self-reported impulsivity scores in methylphenidate-treated participants with ADHD compared with unmedicated participants with ADHD, as well as lower impulsivity on objective behavioural measures (namely shorter SSRT and flatter delay discounting, as indexed by a larger AUC), and c) higher scores in self-reported impulsivity in methylphenidate-treated participants with ADHD compared to healthy controls as well as more pronounced impulsivity in objective behavioural measures (namely longer SSRT and steeper delay discounting as indexed by a smaller AUC).
5. Correlational associations: Based on the above, we expect the following correlations: a) We expect CB1 receptor availability in the dorsal striatum to negatively correlate with motor impulsivity measures; b) We expect CB1 receptor availability in the ventral striatum to be negatively correlated with delay discounting; c) We expect a negative correlation between the availability of CB1 receptors in both the dorsal and ventral striatum and the concentrations of the endocannabinoids AEA and 2-AG in blood plasma; d) We expect a positive correlation of the endocannabinoids AEA and 2-AG in blood plasma with motor impulsivity and delay discounting.

We classify the following questions, for which there is little evidence in the literature, as exploratory in order to generate new hypotheses for future research projects: (1) Does the availability of CB1 receptors differ in frontal brain regions (dorsolateral and ventrolateral cortex)? (2) Do the concentrations of the endocannabinoid-like molecule PEA and the lipids AA and OEA differ in blood serum? (3) Are potential changes in the endocannabinoid system (CB1 receptor availability, concentration of the endocannabinoids AEA and 2-AG in blood plasma) associated with changes in intrinsic network connectivity in ADHD? These exploratory questions relate to the following comparisons: a) unmedicated participants with ADHD versus healthy controls, b) Methylphenidate-treated participants with ADHD compared with unmedicated participants with ADHD, c) Methylphenidate-treated participants with ADHD compared with healthy controls.

## Trial design

In this monocentric, parallel-group study with three study arms, all participants (i.e., unmedicated participants with ADHD, methylphenidate-treated participants with ADHD, and healthy controls) will receive the radiotracer [^18^F]MK-9470. All study participants will undergo the same experimental procedure (Figure 1).

**Figure 1:**
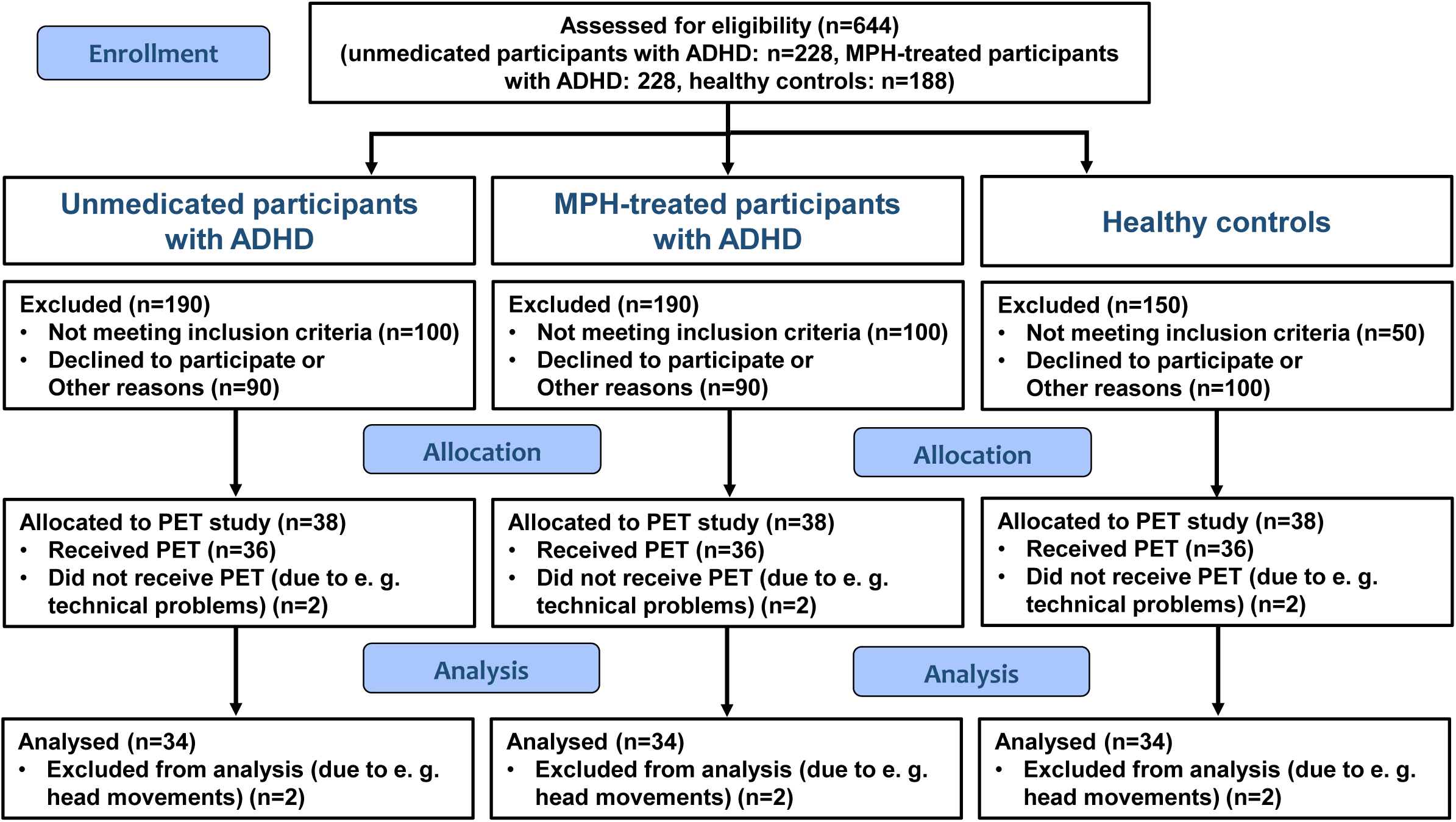
Flow diagram. This is a monocentric, parallel-group study with three study arms (i.e., unmedicated participants with ADHD, methylphenidate-treated participants with ADHD, and healthy controls).

## Methods: Participants, interventions and outcomes

### Study setting

The participating site is the University Medical Center of the Johannes Gutenberg University. The following departments are involved:

Department of Nuclear Medicine, University Medical Center of the Johannes Gutenberg University Mainz, Germany
Department of Psychiatry and Psychotherapy, University Medical Center of the Johannes Gutenberg University Mainz, Germany
Neuroimaging Center, University Medical Center of the Johannes Gutenberg University Mainz, Germany

### Eligibility criteria

#### *Inclusion criteria* for all study participants

Age between 21 and 40 years; fulfillment of all ethical and legal requirements as specified in the study protocol.

Additional *inclusion criteria* for study participants with ADHD:

- DSM-5 diagnosis for ADHD, predominantly hyperactive-impulsive or combined presentation
- Age of onset in childhood
- Group 1 (unmedicated): Off all stimulant medication for at least 6 months, including methylphenidate, amphetamine and lisdexamfetamine, atomoxetine and guanfacine (for medications only approved for use in children and adolescents: off-label use)
- Group 2 (methylphenidate-treated): Stable treatment with methylphenidate for at least the last 3 months (monotherapy)

#### *Exclusion criteria* for all study participants

- History of head injury resulting in brain damage
- History of neurological, endocrine or cardiovascular disorders
- Current or recent (within the last six months) ICD-10 diagnosis of F0x, F1x, F2x, F3x, F4x, F5x
- History of ICD-10 diagnoses F0x, F2x, F3x, F6x, F8x
- Use of all psychotropic drugs within the last 6 months except methylphenidate for participants with ADHD in group 2
- Current or recent (within the last month) recreational use of illicit substances according to the Heidelberg Drug Questionnaire
- Recent (within the last three months) THC use, determined by hair analyses (0.02 ng/mg cut off)
- Positive result in a THC urine test that can detect the THC metabolite THC-COOH for up to 30 days (18 ng/ml cut off)
- Positive result in a multi-field drug test to detect the following substances: Morphine/heroin (opiates) (300 ng/ml cut off), amphetamines (300 ng/ml cut off), cocaine (300 ng/ml cut off), methamphetamines (300 ng/ml cut off), benzodiazepines (100 ng/ml cut off), barbiturates (300 ng/ml cut off), tricyclic antidepressants (300 ng/ml cut off), methadone (300 ng/ml cut off), MDMA/XTC (ecstasy) (500 ng/ml cut off)
- Pregnancy or breastfeeding
- Contraindications to the safety of MRI, including the presence of metal plates, pins, bridges or dentures

### Who will take informed consent?

Informed consent will be obtained by a nuclear medicine physician in accordance with the the Declaration of Helsinki. The written informed consent form, the study protocol and all other written information provided to the subjects were approved by the responsible independent ethics committee.

### Additional consent provisions for collection and use of participant data and biological specimens

Participant data will be handled in accordance with the current Good Clinical Practice Guideline and the Declaration of Helsinki. All biological samples (hair, urine, and blood samples) will be stored in pseudonymized form in secured rooms at the University Medical Center of the Johannes Gutenberg University Mainz. Blood samples will be stored in freezers at -80°C. A separate consent form will be required for analyses of genetic variant from blood samples.

## Interventions

### Explanation for the choice of comparators

Control group participants are aged 21 and 40 years and have no history of ADHD diagnosis. The groups will be matched for age, sex, and intelligence.

### Intervention description

All study participants will undergo identical experimental procedure. The timing and content of study visits are outlined in Table 1. The experimental protocol can be divided into three appointments taking place at the University Medical Center Mainz. These appointments are scheduled approximately 2 weeks apart. Informed consent will be obtained at the first appointment.

**Table 1:** Participant timeline

| TIMEPOINT | STUDY PERIOD |  |  |  |  |
| --- | --- | --- | --- | --- | --- |
|  | Phone Screening | Information and Consent | Assessment Appointments |  |  |
|  | -8 - 0 weeks | 0 week | 0 - 2 weeks | A<sub>1</sub>: 2 - 4 weeks | A<sub>2</sub>: A<sub>1</sub> + 2 weeks |
| **ENROLLMENT:** |  |  |  |  |  |
| Eligibility screen | X |  |  |  |  |
| Informed consent |  | X |  |  |  |
| **ONLINE ASSESSMENTS:** |  |  | X |  |  |
| *WURS-k* |  |  | X |  |  |
| *ADHD-SR* |  |  | X |  |  |
| *CAARS-S:L* |  |  | X |  |  |
| *WRAADDs* |  |  | X |  |  |
| *WST* |  |  | X |  |  |
| *BIS-11* |  |  | X |  |  |
| *UPPS* |  |  | X |  |  |
| *AUDIT* |  |  | X |  |  |
| *FTND* |  |  | X |  |  |
| *HDB* |  |  | X |  |  |
| Drug Screening: |  |  |  | X |  |
| MRI: |  |  |  | X |  |
| **PSYCH. ASSESSMENTS:** |  |  |  | X |  |
| *QB* |  |  |  | X |  |
| *MINI* |  |  |  | X |  |
| *IDA-R* |  |  |  | X |  |
| **BEHAVIORAL ASSESSMENTS:** |  |  |  | X |  |
| *Delay Discounting Task* |  |  |  | X |  |
| *Stop-signal Task* |  |  |  | X |  |
| Blood sampling: |  |  |  |  | X |
| PET: |  |  |  |  | X |

Prior to the second appointment, ten questionnaires in their German versions are to be completed at home on a computer using the online survey software LimeSurvey provided by the University Medical Center Mainz. These comprise questionnaires assessing current and childhood ADHD symptoms (ADHD Self-Assessment Scale (ADHD-SR; (60)); Conner Adult ADHD Rating Scale in the self-long version (CAARS-S:L) (61); Wender-Reimherr Self-Assessment Questionnaire (WRAADDS) (62); Wender-Utah-Rating Scale Short Form (WURS-k; (63)), questionnaires assessing self-rated trait impulsivity (Barratt Impulsiveness Scale-11 (BIS-11) (58); UPPS Impulsive Behavior Scale; (64)), questionnaires assessing substance consumption (Heidelberger Drogenbögen (HDB) (65) as well the following variables: lifetime use (yes/no), age of first use (years), quantity of lifetime use (joints) and time since last use (days); AUDIT: The Alcohol Use Disorders Identification Test (66) as well the variable: duration (years); Fagerström Test for Cigarette Dependence (67)), and the multiple choice vocabulary intelligence test (MWT-B; (68)). At the second appointment, a structural magnetic resonance imaging (MRI) scan of the brain, a resting-state functional MRI (rs-fMRI) measurement, and a psychological examination will be carried out at the University Medical Center Mainz. This examination includes the Mini-International Neuropsychiatric Interview (M.I.N.I) (69), a semi-standardised interview for an integrated diagnosis of ADHD in adulthood (IDA-R interview (70)), the computerised test procedure for objective measurement of the parameters hyperactivity, attention, and impulsivity (Qb-Test; (71)). In order to experimentally assess impulse control capacity, all participants complete the stop-signal and the delay discounting paradigms. In addition, a hair sample will also be taken at this appointment and a rapid drug test will be carried out. If the test is positive, the participant will be excluded from the study.

At the third appointment, blood samples and a PET measurement with the CB1 receptor ligand [^18^F]MK-9470 will be taken at the Department of Nuclear Medicine. From the blood samples, the concentrations of the endocannabinoids AEA and 2-AG, and the compounds AA, PEA and OEA will be determined. Due to the previous findings regarding polymorphisms in the CNR1 gene in connection with impulsivity, the CNR1 polymorphisms from this first blood sample will also be analysed. This will allow to stratify the sample in order to analyse the influence of the genetic effect on our target parameters and to statistically control for these potential effects.

### Criteria for discontinuing or modifying allocated interventions

Given study participants will not be examined further if they withdraw their consent to participate in the study or if their inclusion criteria change during the course of the study and they no longer meet the study criteria (e.g. due to new medication, pregnancy, etc.).

### Strategies to improve adherence to interventions

To improve compliance with the study protocol, participants will be fully informed about each examination. Before to the third appointment (i.e., the PET scan), participants will be contacted by telephone to allow for further questions.

### Relevant concomitant care permitted or prohibited during the trial

The use of tetrahydrocannabinol and other substances that interact with the endocannabinoid system is not permitted during the trial. Substance abuse will be assessed by means of drug testing of hair and urine samples and participants will be informed in advance.

### Provisions for post-trial care

To date, no side effects, complications or consequential damage have been reported as a result of PET scans with [^18^F]MK-9470 or comparable PET tracers. The state of Rhineland-Palatinate provides insurance cover for any health problems that may occur as a result of the examination. In addition, the state of Rhineland-Palatinate provides insurance cover for the use of ionising radiation on humans as part of research projects.

### Outcomes

#### Primary outcome

The primary outcome is the distribution volume V_T_ in the striatum, calculated from the radioactivity concentration in blood and tissue using a two-tissue compartment model (72). It is defined as the ratio of radioligand concentration in tissue to that in plasma (73).

#### Secondary outcomes

Secondary outcomes include:

- Distribution volume V_T_ for cortico-striatal brain regions
- Blood plasma concentrations of the endocannabinoids AEA and 2-AG, and the compounds AA, PEA and OEA
- Parameters from WURS-k, ADHS-SB, Qb-Test, CAARS-S:L and WR-SB for ADHD groups and controls and from IDA-R for ADHD groups
- Parameters from BIS-11 and UPPS
- Motor impulsivity parameters from the stop-signal paradigm (i.e., stop-signal reaction time, trigger failures)
- Delay discounting parameters from the delay discounting paradigm (i.e., discounting rate (*k*) and area under the curve)
- Parameters from intrinsic brain connectivity measures in the resting state fMRI data

#### Outcome measures

Distribution volume V_T_ was chosen as the primary outcome measure, because this parameter achieves a higher sensitivity and specificity in detecting subtle changes in physiological processes as compared to semi-quantitative measures such as the standardized uptake value. The striatum was chosen as a target region because striatal alterations have been demonstrated in ADHD (33). Furthermore, treatment with stimulants that act on the dopamine system has been shown to have therapeutic effects on central dysfunctions in ADHD, namely motor impulsivity and delay discounting (34–38), which are mediated via the ventral and dorsal striatum (8).

Secondary outcomes include distribution volumes in other brain regions relevant to ADHD, endogenous ligands of the endocannabinoid system and related compounds. In addition, behavioural parameters and neuropsychological measures will be collected to assess ADHD symptoms and impulsivity.

### Participant timeline

Screening for eligibility will take place at the outpatient clinic. As part of the study, three appointments will take place at the University Medical Center Mainz. Informed consent will be obtained at the first appointment. Prior to the second appointment, ten questionnaires are to be completed at home on a computer using online survey software provided by the University Medical Center Mainz. This will take approximately 1.5 – 2 hours in total. At the second appointment, structural MRI and rs-fMRI and a neuropsychological assessment will be carried out at the Neuroimaging Center or at the Department of Psychiatry and Psychotherapy. A hair sample will also be taken at this appointment and a rapid drug test will be carried out. If the test is positive, the participant will no longer be able to participate in the study. The entire procedure at this appointment will take approximately 3.25 - 3.75 hours. At the third appointment, blood samples and a PET scan with the tracer [^18^F]MK-9470 will be performed in the Nuclear Medicine Department. The whole procedure on this day will take about 7 hours. In total, the procedure will take about 13 hours (Table 1).

### Sample size

With a two-sided significance level (α = 0.05) and 34 participants with ADHD and 34 healthy controls, this study shows an effect size of 0.7 with 80% power. The third group should also consist of 34 participants to show a similar effect size. The effect size used in the design was estimated from the effect size of the standardised uptake value (SUV) from previous publications (40,47,74) using G*Power 3.1. The publications had an average effect size of 0.95. As the published studies had very small case numbers, we chose a more conservative effect size estimate of 0.7 (range: 0.66 - 1.47). Power planning using ANCOVA may result in a lower number of participants with ADHD or healthy controls required, depending on the assumed correlation of potential confounders (see below) with the outcome variable V_T_. We have no prior knowledge of the correlation between the potential confounders and the outcome variable V_T_. Therefore, the sample size was calculated using a t-test, although this approach is conservative. The sample size is increased by 10 % to 38 per group to account for potential drop-outs.

### Recruitment

Participants with ADHD will be recruited from the outpatient clinic for Psychiatry and Psychotherapy at the University Medical Center Mainz. The ADHD outpatient clinic has already been successfully involved in several studies in adult ADHD, including the BMBF funded ESCAlife program. Approximately 120 individuals with ADHD per year are seen at the outpatient clinic for diagnosis and treatment. Experience has shown that outpatients are intrinsically very interested in participating in scientific studies. We therefore expect a high level of willingness to participate on the part from this clientele. In addition, we informed the ADS-Mainz e. V. association (Wilhelm-Busch-Straße 20, 55126 Mainz) about the planned study and a presentation and discussion of the study is planned. The ADS-Mainz e. V. association works with individuals with ADHD, parents, educators and teachers as well as other interested parties and promotes further education in this field. The ADS-Mainz e. V. association has already indicated its willingness to support the recruitment of individuals with ADHD and healthy controls. Healthy controls will be recruited via notices at the University Medical Center and the Johannes Gutenberg-University Mainz, as well as through student classes at the Department of Nuclear Medicine.

## Assignment of interventions: allocation

### Sequence generation

n/a, as all study participants will undergo identical experimental procedures. Group assignment is based on diagnosis (ADHD vs. No ADHD) and medication status (methylphenidate-treated vs. unmedicated).

### Concealment mechanism

n/a, as “Sequence generation” is not applicable.

### Implementation

n/a, as “Sequence generation” is not applicable.

## Assignment of interventions: Blinding

### Who will be blinded

n/a, as all study participants will undergo identical experimental procedures.

### Procedure for unblinding if needed

n/a, as “Blinding” is not applicable.

## Data collection and management

### Plans for assessment and collection of outcomes

The data of the study participants will be assessed and collected by the teams of the Department of Nuclear Medicine and the Department of Psychiatry and Psychotherapy and entered into a table designed for this study. The principal investigator or designated representatives are responsible for ensuring that the table is correctly completed and that entries can be verified against source data. The principal investigator or designated representatives should complete the table entries as soon as possible after the information is collected. Entries in the table are checked using the dual control principle. Any outstanding entries must be completed immediately after the final examination. An explanation should be given for any missing data. Diagnostic and clinical interviews and examinations will be carried out by clinical psychologists or appropriately trained staff. The data quality of the MRI datasets will be assessed using state-of-the-art methods (75).

Below we describe the questionnaires used in our study, along with their reliability and validity. The Wender Utah Rating Scale-Short Version (WURS-k) is a self-report instrument for the retrospective assessment of childhood ADHD symptoms in adults. It includes 25 items scored on a five-point scale. The instrument shows good split-half reliability (r12=0.85) and strong internal consistency (α=0.91), as well as a good ability to distinguish between individuals with and without childhood ADHD (63).

The ADHD Self-Rating Scale (ADHS-SR) is a tool for measuring current ADHD symptoms in adults according to diagnostic criteria. It consists of 18 items that are rated on a four-point scale. The test shows good test-retest reliability (0.80) and internal consistency (0.90) and an acceptable ability to distinguish between those with and without ADHD at various cut-off values (76).

The Conners’ Adult ADHD Rating Scales-Self-Report: Long Version (CAARS-S:L) is a tool for measuring ADHD symptoms in adults (77). It comprises 66 items rated on a four-point scale. Depending on the subscale, the test shows acceptable to excellent internal consistency and good to excellent test-retest reliability, as well as high sensitivity and specificity (78,79).

The Wender-Reimherr Adult Attention Deficit Disorder Scale (WRAADDS) is an instrument for assessing ADHD symptoms in adults. It comprises 59 items that are rated on a five-point scale. The 10 subscales of the instrument are proven to have good to excellent internal consistency as well as strong validity through correlations to ADHD symptom checklists (80). The Vocabulary Test (WST), an intelligence test developed by Schmidt & Metzler in 1992, has become established as a quick way of measuring intelligence. The test comprises 42 questions in which a real word must be selected from a list of similarly written but invented words. The test has been shown to correlate with the HAWIE-R intelligence test, but to overestimate it somewhat (81). The split-half reliability is r = 0.95 and the internal consistency α = 0.94 (82).

The Barratt Impulsiveness Scale-11 (BIS-11) is a self-report instrument for measuring impulsivity. It consists of 30 items scored on a four-point scale. The BIS-11 captures six subscales of impulsivity, including attention, motor, self-control, cognitive complexity, perseverance, and cognitive instability. The test has acceptable internal consistency (α = 0.69), with the removal of item 11 (“sliding back and forth”) resulting in a relative improvement to α = 0.74 (83). Reliability measures for the English version of the test, with a test-retest reliability of 0.83 being found (84).

The UPPS Impulsive Behavior Scale is a scale developed to understand impulsivity (85). The scale includes four subscales labeled urgency, (lack of) premeditation, (lack of) perseverance, and sensation seeking. Cronbach’s α for each of the subscales (German version) were reported respectively at 0.82, 0.80, 0.85, and 0.83 (86).

The AUDIT (Alcohol Use Disorders Identification Test) is a screening tool for identifying alcohol use disorders. It consists of 10 questions that are scored on a four-point scale. The AUDIT assesses alcohol consumption behavior and the risk of dependence and harm from alcohol consumption. The AUDIT was assessed for its validity (87) and was found to have an interclass correlation coefficient of 0.95 and showed good sensitivity and specificity for alcohol dependence, alcohol use disorder (AUD), as well as those at risk for AUD.

The FTND (Fagerström Test for Nicotine Dependence) is a self-assessment tool for evaluating nicotine dependence. It consists of six questions covering aspects such as the number of cigarettes smoked per day, the craving for the first morning cigarette and the difficulty of smoking in smoke-free areas. Higher scores on the FTND indicate higher nicotine dependence. The test assesses the severity of dependence and monitors the effectiveness of smoking cessation programs.

The Heidelberg Drug Questionnaire (HDB) is a versatile instrument for assessing the use and risk level of illegal drugs. It is suitable for use with young people aged 12 and over and adults. The HDB is characterized by its modular structure, which enables a differentiated assessment of the use of five substance groups: cannabis, amphetamines, ecstasy, cocaine and hallucinogens. Two modules are available for each of these substance groups: 1) Knowledge module: This module comprises 15 items and records the respondent’s knowledge of the respective drug, including its mode of action, consumption risks and side effects. 2) Behavior module: This module comprises 16 items and records the respondent’s individual consumption pattern in the last 12 months. This includes the dosage, frequency of use, mixed use with other drugs, motives for use and the chosen contexts of use. A summary of data management and confidentiality was included in both the study protocol and the informed consent form given to the study participants. The teams are trained before the start of the study to ensure correct data entry. All modules are reported to have acceptable to good internal consistencies, for knowledge modules between α = 0.62 (cannabis knowledge module) and α = 0.90 (amphetamine knowledge module) and behavioral modules α = 0.77 (hallucinogens behavior module) and α = 0.89 (cannabis behavior module) (65).

For PET measurement, [^18^F]MK-9470 will be synthesised as previously described (88). It will be administered as a single intravenous bolus injection; the average injected radioactivity will be 250 MBq in a volume of 10 to 20 ml. PET data will be acquired using a Siemens Biograph Vision PET/CT scanner with an axial field of view of 26.3 cm and a transverse spatial resolution of 3.6 mm at 1 cm (89). Participants will be placed in head first supine position. Prior to performing the PET acquisition blocks, a low-dose CT scan will be performed to allow for attenuation correction. The PET data will be acquired in three parts: The first part will last 90 minutes, after a break of 30 minutes, the second part of the measurement will last 60 minutes. After 360 minutes, a late measurement will take place in order to assess the entire time course of the tracer. If, after a methodical subgroup analysis, it turns out that this measured value does not provide any added value for the evaluation, it will not further recorded in order to shorten the protocol. The data will be reconstructed with the Poisson ordered subset expectation maximization algorithm using time-of-flight and point spread function with 4 iterations and 5 subsets, resulting in an image matrix of 440 × 440 × 75 and voxels of 0.8 × 0.8 × 1.5 mm^3^. Further key performance characteristics of the PET/CT-scanner are described in the publication by Reddin et al. (89). Motion artefacts in PET images are corrected using the realignment algorithm implemented in Statistical Parametric Mapping (SPM) 12 (90). The correction of partial volume effects will be performed with the SPM12 integrated toolbox "PETPVE12". As a prerequisite, PET images are coregistered with corresponding structural MR images using the coregistration algorithm implemented in SPM12. The segmentation of the MRI data into grey matter, white matter and cerebrospinal fluid will be covered by the "PETPVE12" toolbox, which accesses the algorithm of the "VBM8" toolbox integrated in SPM12.

Parameter calculation will be based on pharmacokinetic modelling using a reversible two-tissue compartment model (72) using the PMOD software (PMOD Technologies LLC). The time course of the arterial radioactivity concentration required for the model will be quantified on an image basis by segmenting the carotid artery. Blood samples are taken to determine and correct the amount of metabolites in the signal. In a methodological subproject, multilinear analysis methods (91) are evaluated for the analysis of [^18^F]MK-9470, which has not yet been done for this ligand, as provided by the PMOD software. Furthermore, MR images are used for stereotactic normalisation using the DARTEL toolbox as implemented in SPM12.

The endocannabinoids AEA and 2-AG and the compounds AA, PEA and OEA are extracted from 500-µl blood plasma samples and determined according to in-house protocols (92,93). In addition, CNR1 polymorphisms will be analysed from 5-ml blood plasma samples and determined according to a published protocol (94).

### Plans to promote participant retention and complete follow-up

Experience has shown that outpatients are very interested in participating in scientific studies. This also applies to the study participants who form the control group. Therefore, a high level of study compliance and a low drop-out rate can be expected. To minimize the risk of participant retention, the study staff provides close and personal support to participants, e.g., personal and telephone contact during study participation.

### Data management

Study data will be collected electronically using a table designed for this study. Upon completion of data entry, all data will be locked, access to data entry will be blocked, and data will be checked for plausibility, consistency, and completeness using the four-eye principle. If no further corrections are required, the table will be declared closed and used for statistical analysis. The data checks are carried out by the P.I.s or a designated representative at the University Medical Center Mainz. Only pseudonymized data will be used for data extraction, analysis and dissemination. The Department of Nuclear Medicine and the Department of Psychiatry and Psychotherapy will be responsible for data management and archiving. Data will be stored in pseudonymized form for 30 years.

### Confidentiality

Electronic data will be kept strictly confidential and inaccessible to any unrelated individuals or third parties throughout the trial. Participants’ names and other confidential information will be protected by medical confidentiality and will not be disclosed to the sponsor. During the study, each participant will be identified only by a unique identification code. A personal subject identification list (linking participant codes to corresponding names) will be securely maintained by the investigator to allow for record identification. The teams in each participating department will be responsible for ensuring confidentiality, adherence to the Good Clinical Practice guidelines and compliance with local data protection laws. To further secure data, organisational procedures will prevent unauthorised access.

All data management procedures at the center will be carried out according to written standard operating procedures (SOPs) of the Department of Nuclear Medicine and the Department of Psychiatry and Psychotherapy, which will ensure efficient conduct in accordance with good clinical practice (GCP) and in strict compliance with national and EU regulations. Upon study completion, the data will be converted into archival data formats for archiving to ensure their reuse.

### Plans for collection, laboratory evaluation and storage of biological specimens for genetic or molecular analysis in this trial/future use

All biological samples (hair, urine, and blood samples) will be stored in pseudonymized form in secured rooms at the University Medical Center of the Johannes Gutenberg University Mainz. Blood samples will be stored in freezers at -80°C. Only pseudonymised biological samples will be forwarded for further analysis, e.g., to the Institute of Forensic Medicine or Human Genetics at the University Medical Center Mainz.

## Statistical methods

### Statistical methods for primary and secondary outcomes

The primary analysis will be the group comparison (unmedicated participants with ADHD vs. healthy controls) of the volume of distribution V_T_. This will be done using an analysis of covariance (ANCOVA). Fixed effects are "group" and "sex". "Age” is considered as a random effect. It has been shown that the distribution of cannabinoid type 1 receptors changes with age and shows differences between men and women (95). If the V_T_ is not normally distributed, a monotone transformation of V_T_ will be considered. The analysis will be checked using residual plots. The two-sided significance level is set at 5 %. The primary analysis population consists of all enrolled participants with a valid PET and existing fixed and random effects. Models with the additional fixed effects of current alcohol use, current tobacco use, previous cannabis use and their interaction with group, if applicable, are used as sensitivity analyses. Tobacco, cannabis and alcohol use have been shown to reduce the availability of CB1 receptors in the brain (39,41,42). All secondary and exploratory variables are compared between groups using ANOVA/ANCOVA methods. All primary and secondary parameters will be presented descriptively and with effect sizes where appropriate. The analysis will be pre-specified in a statistical analysis plan.

### Interim analyses

n/a; there are no interim analyses.

### Methods for additional analyses (e.g. subgroup analyses)

Depending on the sample, additional variables may be added to the statistical model to adjust for other possible confounders.

### Methods in analysis to handle protocol non-adherence and any statistical methods to handle missing data

It is assumed that data may be missing mainly due to technical failures. Such missing data are treated as missing completely at random (MCAR) and missing values are not imputed.

### Plans to give access to the full protocol, participant level-data and statistical code

The study has been registered in a national register The study protocol will be available upon request after publication of the results. The study dataset will remain confidential until the final analysis has been performed and published. According to a recent consensus statement on best practices to share and reuse data from clinical trials, de-identified individual participant data will be available in a suitable data repository on request after the end of this proposed study (e. g., for meta-analyses).

## Oversight and monitoring

### Composition of the coordinating centre and trial steering committee

Coordinating Center: The study will be coordinated by the Department of Nuclear Medicine, University Medical Center of the Johannes Gutenberg University Mainz, Germany, with IM [PI MIEDERER] serving as the coordinating investigator. The center is responsible for overall study management, data coordination, and regulatory compliance.

Trial Steering Committee: The committee comprises: Principal Investigator and PET imaging specialist [IM], Co- Principal Investigator [AS].

The committee will meet at least monthly during the first three months, then quarterly thereafter, with additional meetings as needed. Responsibilities include: Monitoring study progress and safety, reviewing protocol deviations, ensuring adherence to Good Clinical Practice, making decisions on study modifications, overseeing data quality and integrity.

Study Team: The operational study team will meet weekly throughout the study duration to discuss: Patient recruitment and scheduling, protocol adherence and safety monitoring, data collection and quality assurance, operational issues and problem resolution, coordination between different study components (PET imaging, clinical assessments, laboratory analyses).

### Composition of the data monitoring committee, its role and reporting structure

A formal data monitoring committee (DMC) is not planned for this study. This decision is justified by the following factors: Single-center, investigator-initiated study with limited sample size, use of established, well-characterized PET tracer [^18^F]MK-9470, no therapeutic intervention beyond standard clinical care, no vulnerable populations (participants ≥18 years, capable of informed consent), low risk of serious adverse events related to study procedures, safety monitoring will be conducted by the trial steering committee through regular review of adverse events and protocol deviations. The study can be terminated at any time by the principal investigators if safety concerns arise.

### Adverse event reporting and harms

Documentation of any unintended effects in relation to the study and initiation of appropriate action will be in place.

### Frequency and plans for auditing trial conduct

To determine how the study is progressing and to identify any changes that need to be made, the recruitment status of the study is reviewed on a monthly basis.

### Plans for communicating important protocol amendments to relevant parties (e.g. trial participants, ethical committees)

Any major changes to the protocol that affect the conduct of the study, including changes to the study design, sample size, procedures, or objectives, will require a formal amendment to the study protocol. Formal amendments to the protocol will be formally submitted to the responsible local Ethics Committee. Major amendments to the protocol will only be implemented after approval by the Ethics Committee.

## Dissemination plans

New therapeutic approaches for ADHD are urgently needed. The study is registered in an international registry. International guidelines such as SPIRIT (96) and CONSORT (97) will be followed. The results will be presented at national and international meetings and published in an international peer-reviewed journal.

## Discussion

New treatments are urgently needed to better address the clinical needs of individuals with ADHD. The stimulants methylphenidate and amphetamine, the first-line drugs, are very effective. Unfortunately, not all individuals with ADHD respond to them and in some cases their use is contraindicated. In addition, they do not meet the wider clinical needs of individuals with ADHD, especially in the longer term. A new and promising approach to better understand the underlying neural mechanisms of ADHD and to develop new therapeutic approaches is to investigate the involvement of the endocannabinoid system. Due to its ubiquitous distribution in the brain and modulation of neurotransmitter release, the endocannabinoid system is increasingly implicated in mental disorders, including ADHD (26). ADHD has previously been linked to the dopamine system: striatal alterations have been demonstrated in ADHD (33), and stimulant treatment that acts on the dopamine system has been shown to have therapeutic effects on central dysfunctions in ADHD, namely motor impulsivity and delay discounting (34–38). Importantly, the dopamine system is modulated by the endocannabinoid system (11,23,24) and there is evidence for the reverse (25). Given the dopamine hypothesis, the dopaminergic involvement in the striatum and in motor impulsivity and delay discounting in ADHD, and the interaction between the dopamine and endocannabinoid systems, it is conceivable that the endocannabinoid system may be involved in the aetiology and pathogenesis of ADHD. The aim and novelty of this basic research-oriented study is therefore to investigate, for the first time to our knowledge, the role of the striatal endocannabinoid system in medication-free and methylphenidate-treated participants with ADHD using [^18^F]MK-9470 PET. In secondary analyses, based on the predictions of the "dual pathway model of ADHD", we will investigate the availability of CB1 receptors in both the dorsal and ventral striatum. We will also assess circulating endocannabinoid levels and impulsive measures related to motor impulsivity and delay discounting. We will also assess correlations between these modalities. The present study aims to elucidate novel aspects underlying the pathophysiology of ADHD as a starting point for the development of new pharmacological treatment approaches for ADHD, which are currently lacking. Questions related to the physiology and pathophysiology of the endocannabinoid system are highly relevant to the current discussion on the legalisation of cannabis, the use of cannabis in medicine in general and the self-medication of individuals with ADHD in Germany. Therefore, our study will make a significant contribution to the understanding of this area.

CB1 receptors are visualised with the radioactive ligand [^18^F]MK-9470. This radioligand acts as an inverse agonist that binds to the cannabinoid type 1 receptors with high selectivity and affinity (human IC_50_ = 0.7 nM), is very lipophilic (logD7.3 = 4.7) and shows good uptake in the brain (20,98). The radioligand [^18^F]MK-9470 was selected because it has comparable in vitro and in vivo properties to the successfully used radioligand [^18^F]FMPEP-d2 (90), but can be prepared in a one-step procedure by direct ^18^F labelling instead of a two-step synthesis and achieves a radiochemical yield of 30.3 ± 11.7 % (88).

[^18^F]MK-9470 PET data can be analysed in various ways, including semi-quantitative methods, with the most sophisticated approach being the use of mathematical models. The application of a model requires the time course of the radioactivity concentration in the arterial blood and tissue. Together with a known model configuration, the model parameters describing the uptake and binding of the radiotracer in the tissue can be estimated (for detailed information see (99)). Tracer kinetic modelling allows a precise quantification of the processes in the tissue beyond a simple uptake measurement. Therefore, the parameter calculation for [^18^F]MK-9470 will be based on the use of a reversible two-tissue compartment model (72). As [^18^F]MK-9470 has slow tracer kinetics, a 180 min acquisition protocol with a late time point at 360 min will be required to cover the full kinetics. To quantify the radioactivity concentration in the blood, the image data of the carotid artery will be segmented and corrected for metabolites. In a methodological subproject, multilinear analysis methods (91) will be investigated for the analysis of [^18^F]MK-9470, which has not yet been done for this ligand. This approach is pursued as linearisation reduces the sensitivity to image noise and allows more robust and stable estimates of kinetic parameters. This novel and unique basic research approach will provide important insights into the endocannabinoid system and potential alterations in adult ADHD. In particular, the CB1-ADHD study will contribute to a better understanding of striatal CB1 receptor availability and the association of alterations in the endocannabinoid system with various aspects of impulsivity present in ADHD. The CB1-ADHD study will include an unusually large sample for a PET study (i.e., 38 participants per group). However, the results need to be replicated in follow-up studies. Nevertheless, the results will provide an important starting point for the development of new pharmacological treatments for ADHD, which are currently lacking. The results of this study will therefore have a major impact on future basic and clinical research and will help to open up new avenues for the development of evidence-based therapeutic strategies for the treatment of ADHD.

## Trial status

The currently used version of the study protocol is version 6, 5 July 2025. The recruitment for this study started on 1 October 2025. We expect to recruit the required 114 study participants within 2 years.

## Abbreviations

AA: arachidonic acid
ADHD: Attention deficit/hyperactivity disorder
AEA: N-arachidonoylethanolamine (anandamide)
AUC: area under curve
CB1: cannabinoid type 1
OEA: oleoylethanolamide
PEA: palmitoylethanolamide
PET: positron emission tomography SSRT stop-signal reaction time 2-AG 2-arachidonoylglycerol

## Data Availability

All data produced in the present study are available upon reasonable request to the authors.

## Acknowledgements

n/a; no one contributed towards the article who does not meet the criteria for authorship.

## Authors’ contributions

IM and AS conceived the study idea and acquired funding. IM and AS initiated the study design and will provide overall project supervision. IM will serve as principal investigator and oversee project administration together with AS. MLS will be responsible for data collection and investigation procedures. CR provided statistical expertise in study design and power analysis and will supervise the planned statistical analyses. WR, and MS provided essential resources for study implementation. IM drafted the study protocol and all authors contributed to protocol refinement and approved the final version. IM and AS will ensure execution and completion of the project, including data curation and methodology oversight. IM and AS will draft manuscripts for publication arising from this study with contributions and approval of final versions from all authors.

## Availability of data and materials

Only members of the study group will have access to the final study dataset. The dataset will be kept confidential until the results are published.

## Ethics approval and consent to participate

The study has been approved by the Ethics Committee of the Landesärztekammer Rheinland-Pfalz (Ethics Committee of the District Medical Council of Rhineland Palatinate - approval number 2021-15797). The study was registered in the German Clinical Trial Register (DRKS-ID: DRKS00037526). Informed consent will be obtained from all study participants. All procedures in this study will be performed in accordance with the Declaration of Helsinki.

## Consent for publication

All participants in the study will give their informed consent for the use of their data.

